# National Assessment on the Frequency of Pain Medication Prescribed for Intrauterine Device Insertion Procedures within the Veterans Affairs Health Care System

**DOI:** 10.1101/2024.07.25.24311008

**Authors:** Anna D. Ware, Terri L. Blumke, Peter J. Hoover, Zach P. Veigulis, Jacqueline M. Ferguson, Malvika Pillai, Thomas F. Osborne

## Abstract

**Background:** The intrauterine device (IUD) is a highly effective form of long-acting reversible contraception, widely recognized for its convenience and efficacy. Despite its benefits, many patients report moderate to severe pain during and after their IUD insertion procedure. Furthermore, reports suggest significant variability in pain control medications, including no adequate pain medication. The aim of this evaluation was to assess the pharmaceutical pain medication types, proportions, and trends related to IUD insertion procedures within the Veterans Health Administration (VHA).

**Methods:** IUD insertion procedures documented in the VA electronic health record were assessed from 1/1/2018 to 10/13/2023. Descriptive statistics described patient and facility characteristics while annual trends were assessed using linear regression.

**Results:** Out of the 28,717 procedures captured, only 11.4% had any form of prescribed pain medication identified. Non-Steroidal Anti-Inflammatory Drugs (NSAIDs) were the most frequently prescribed pain medication category (8.3%), with ibuprofen being the most common pain medication overall (6.1%). Over the assessment period, there was an average annual increase of 0.52% (*p*=0.038) of procedures with prescribed pain medication, increasing from 10.3% in 2018 to 13.3% in 2023.

**Conclusions:** Although IUD insertion procedures have been seeing an increase in prescribed pain medication, the overall proportion remains disproportionality low relative to the pain experienced. Additionally, when pain interventions were initiated, they disproportionally utilized medication that have been shown to be ineffective. The intent of the work is that the information will help guide data driven pain medication strategies for patients undergoing IUD insertion procedures within the VHA.

## Introduction

Intrauterine devices (IUDs) are a highly recommended long-acting reversible contraception (LARC) option not only due to their mean one-year failure rate of less than 1%, but also because many patients prefer them for their convenience, minimal maintenance, and long-term effectiveness.^1,2^ However, pain surrounding the insertion of the IUD has been gaining widespread attention due to numerous negative patients experiences.^3,4,5,6^ In a recent survey, 78% of respondents rated their IUD insertion pain as moderate to severe, with 46% experiencing vasovagal symptoms.^5^ In attempts to mitigate pain during IUD insertion, several pharmacological interventions have been tested with mixed results.^6,7,8,9,10,11,12,13^ For example, certain formulations of lidocaine have been effective, while cervical ripening agents (prostaglandins) and most non-steroidal anti-inflammatory drugs (NSAIDs) were ineffective in managing pain during or after insertion.^9–11^

During the procedure, the IUD is inserted by clamping the cervix using a tenaculum, followed by measuring the depth of the uterine cavity, placement of the IUD, trimming of the strings, and confirmation of appropriate placement. ^14^ While some healthcare providers refer to the sensation caused by the cervical clamping as a ‘quick pinch’, many patients have reported severe pain. ^5,15^ The cervical clamping is often then followed by menstrual-pain-like cramps as the IUD is inserted into the uterus. ^16^ For some, the pain of insertion lasts considerably longer than the approximately 15-minute procedure.^16^ As a result, many patients have called for a comprehensive strategy to manage pain associated with IUD insertion within the United States _(US)._^15,17,18,19,20^

The Veterans Health Administration (VHA) is the largest integrated healthcare system in the US, serving over 9 million enrolled Veterans annually.^21^ Women currently represent approximately 10% of enrolled Veterans, a figure that is expected to rise to 18% by 2040. ^22,23^ Recently, the White House released an executive order to announce new actions to advance health research and innovation for women and gender-diverse individuals to “improve women’s lives across America.”^24^ Additionally, to meet the needs of this rapidly expanding population, VHA has focused on key initiatives to enhance access to comprehensive women’s healthcare through specifically trained providers. ^25,26^ Veterans can now access a full range of contraceptive options with no or minimal out-of-pocket expenses as part of their comprehensive primary care. ^27^ Consequently, LARC use, including IUDs and implants, have been reported to be higher among Veterans who are assigned female at birth (AFAB) of childbearing age (18-45 years) than that of the general US population (23% vs. 11%), with 65% of these Veterans receiving their IUD within a VA outpatient clinic.^22,28^

Yet, despite the growing numbers of women Veterans within VHA, and the higher prevalence of LARC use among Veterans compared to the general US population, it is unknown how frequently pain medication is prescribed for IUD insertion procedures. In this assessment, we examined the type and extent to which pain medication is prescribed for IUD insertions performed in an outpatient setting among Veterans receiving care within the VHA. The intent is that this information and knowledge will help guide data driven quality improvement efforts within the VHA.

## Materials and Methods

### Data Sources

We performed a retrospective assessment of electronic health record (EHR) data for patients who were AFAB, aged 18 years or older, and were receiving care at VA medical centers. The assessment focused on those who underwent an IUD insertion procedure within an outpatient clinic or outpatient surgical visit at a VHA facility from January 1^st^, 2018, to October 13^th^, 2023. Procedures were captured using the CPT code 58300: “Insertion of IUD.”^29^ Outpatient visits where other procedures were performed other than just the IUD insertion (for example, the visit included CPT codes 58300: “Insertion of IUD” and 56740: “Excision of Bartholin’s gland or cyst”) were excluded from our assessment as it could not be determined if prescribed pain interventions were intended for the purpose of the IUD insertion or the other procedures present. All data was queried from the VA’s Corporate Data Warehouse (CDW), which is a relational database that aggregates EHR data from all VHA facilities. ^30^ Prescribed non-opioid analgesic pain interventions were queried from outpatient medication tables at the IUD consult or within (-) 45 days, (+) 1 day surrounding the procedure, as preprocedural consults typically occur within 45 days prior to procedure. Any prescribed opioid analgesics, a highly monitored drug class within VHA, were captured within (+/-) 24 hours of the procedure. Lastly, leveraging SQL string search techniques, we parsed nursing text orders and patient discharge instructions within procedure encounter notes to capture any medications not already documented in structured data within (+/-) 24 hours surrounding the procedure. This allowed the inclusion of pain medications prescribed and recorded through both structured and unstructured EHR data.

### Pharmacological IUD Pain Medication Interventions

Prescribed pharmacological pain interventions associated with the IUD insertion procedure were grouped into five categories, as documented elsewhere:^6,7^ (1) NSAIDs, (2) Lidocaine, (3) Prostaglandins, (4) Opioid Analgesics, and (5) Combination or Other. List of drugs included in each category are available in **Appendix 1**.

### Co-variates

Patient sociodemographic variables included age, self-identified race or ethnicity, marital status, third-party (non-VA) insurance, and rurality of patient’s residence. Additional clinical characteristics included body mass index (BMI), parity status (nulliparous vs. parous), Charlson Comorbidity Index (CCI) score, and female-specific comorbidities (e.g., chronic pelvic pain, postpartum depression, dyspareunia, history of military sexual trauma (MST)). We also captured provider and facility characteristics including the 4 US regions (as defined by the Centers for Disease Control, 2023),^31^ facility complexity (VHA classification system for VA medical centers that ranges from high, medium, and low)^32^, state location of the clinic, outpatient care setting (e.g., gynecology clinic, primary care clinic), and the type of primary provider (e.g., gynecologist, nurse practitioner) associated with the IUD insertion procedure. High complexity facilities have larger levels of patient volume, higher patient risk, more teaching and research resources, and contain level 4 to 5 intensive care units compared to low complexity facilities which have little or no teaching/research, the lowest number of physician specialists per patient, and contain level 1 and 2 intensive care units.^32^

### Statistical Analysis

The data collected for this assessment was analyzed from October 14^th^, 2023, to July 17^th^, 2024. Descriptive statistics to summarize patient, provider, and facility characteristics and compared differences in the prevalence of these characteristics among those prescribed pain medication compared to those without. Differences between groups were evaluated using Chi-square and the student’s t-tests. To quantitatively assess any temporal trends, we applied a linear regression model to evaluate any changes in the number of IUD insertions with prescribed pain medications by year. Data were analyzed using R Statistical Software (v4.1.2; R Core Team 2021). This quality assessment project received determination of non-research from Stanford Institutional Review Board, (Stanford University, Stanford, CA, USA) Protocol #74380.

## Results

### National VHA Results

Among the 1,614,650 patients who were AFAB, aged 18 years or older, and who received care within the VHA during the assessment period, 28,717 IUD insertion procedures were performed across VHA, nationally (flowchart available in **Appendix 2**). Out of the 28,717 procedures captured, 11.4% (3,260) had any form of pharmaceutical pain management prescribed (**Table 1**). When analyzing annual trends, the slope of the fitted linear regression model indicated that, on average, there was a statistically significant annual increase of 0.52% (*p* = 0.038; **Figure 1**) in the percentage of procedures with prescribed pain medication, increasing from 10.3% in 2018 to 13.3% in 2023.

**Figure 1:**
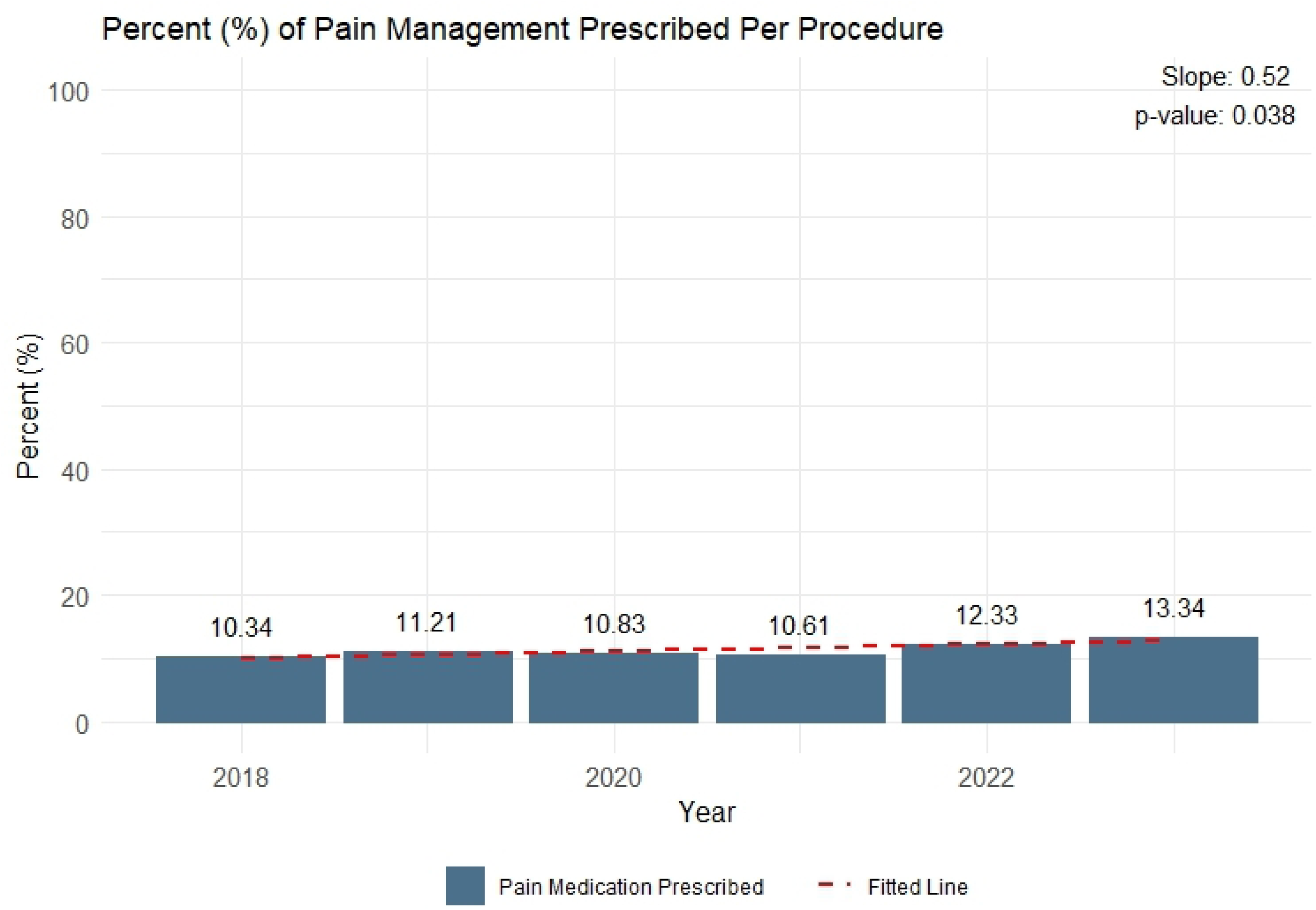
Percent of IUD insertion procedures among the 24,010 IUD insertions performed within VHA between January 1st, 2018, to October 13th, 2023, with any pharmaceutical pain medication prescribed within the Veterans Affairs Healthcare System from January 1^st^, 2018, to October 13^th^, 2023. **F1L:** This graph illustrates the annual percentage of intrauterine device insertion procedures with any pharmaceutical pain medication prescribed within the Veterans Affairs Healthcare System from January 1^st^, 2018, to October 13^th^, 2023. The navy bars represent the mean percentage of procedures with pharmaceutical pain medication prescribed per year. The dashed red line depicts the linear regression line fitted to the observed data. The slope of this line with a p-value indicates the statistical significance of this trend. Each observed data point is labeled with the corresponding percentage.

**Table 1:** Patient characteristics among the 24,010 IUD insertions performed within VHA between January 1st, 2018, to October 13th, 2023, with any documented pain medication for their intrauterine device insertion procedure compared to those without any prescribed pain medication.

|  | Total<br>(Column<br>frequencies) | No Pain<br>Medication<br>Prescribed | Pain<br>Medication<br>Prescribed | *P-value*<sup>a</sup> |
| --- | --- | --- | --- | --- |
| **Total, n (%)** | **28,717 (100.0%)** | **25,457 (88.7%)** | **3,260 (11.4%)** |  |
| ***Sociodemographic characteristics*** |  |  |  |  |
| **Age (years), mean (SD)** | 39.53 (7.58) | 39.58 (7.56) | 39.08 (7.71) | 0.135 |
| **Race and/or Ethnicity, n (%)** |  |  |  | **<0.001** |
| American Indian or Alaskan Native | 412 (1.4%) | 378 (91.7%) | 34 (8.3%) |  |
| Asian | 884 (3.1%) | 773 (87.4%) | 111 (12.6%) |  |
| Black or African American | 7,165 (25.0%) | 6,246 (87.2%) | 919 (12.8%) |  |
| Hispanic | 3,951 (13.8%) | 3,513 (88.9%) | 438 (11.1%) |  |
| Native Hawaiian or Pacific Islander | 340 (1.2%) | 295 (86.8%) | 45 (13.2%) |  |
| Non-Hispanic White | 14,089 (49.1%) | 12,598 (89.4%) | 1,491 (10.6%) |  |
| Declined to Answer or Unknown | 1,876 (6.5%) | 1,654 (88.2%) | 222 (11.8%) |  |
| **Marital Status, n (%)** |  |  |  | **<0.001** |
| Married | 10,649 (37.1%) | 9,574 (89.9%) | 1,075 (10.1%) |  |
| Single | 8,620 (30.0%) | 7,427 (86.2%) | 1,193 (13.8%) |  |
| Divorced/Separated | 8,780 (30.6%) | 7,861 (89.5%) | 919 (10.5%) |  |
| Widowed | 143 (0.5%) | 127 (88.8%) | 16 (11.2%) |  |
| Unknown | 525 (1.8%) | 468 (89.1%) | 57 (10.9%) |  |
| **Has third party (non-VA) insurance, n (%)** |  |  |  | 0.996 |
| No | 19,351 (67.4%) | 17,153 (88.6%) | 2,198 (11.4%) |  |
| Yes | 9,348 (32.6%) | 8,286 (88.6%) | 1,062 (11.4%) |  |
| Unknown | 18 (0.1%) | 18 (100.0%) | - |  |
| **Rurality of Patient's Residence, n (%)** |  |  |  | **0.003** |
| Urban | 22,374 (77.9%) | 19,713 (88.1%) | 2,661 (11.9%) |  |
| Rural | 6,171 (21.5%) | 5,593 (90.6%) | 578 (9.4%) |  |
| Highly Rural | 133 (0.5%) | 117 (88.0%) | 16 (12.0%) |  |
| Unknown | 39 (0.1%) | 34 (87.2%) | 5 (12.8%) |  |
| ***Clinical characteristics*** |  |  |  |  |
| **Body Mass Index, mean (SD)** | 31.30 (6.44) | 31.33 (6.42) | 31.11 (6.59) | 0.798 |
| **Parity Status<sup>b</sup>, n (%)** |  |  |  | **0.002** |
| Nulliparous | 27,114 (94.4%) | 23,997 (88.5%) | 3,117 (11.4%) |  |
| Parous/Multiparous | 1,603 (5.6%) | 1,460 (91.1%) | 143 (8.9%) |  |
| **Charlson Comorbidity Index, mean (SD)** | 0.57 (1.12) | 0.58 (1.13) | 0.55 (1.05) | 0.181 |
| ***Diagnosed Conditions*** |  |  |  |  |
| **Chronic pelvic pain, n (%)** |  |  |  | **<0.001** |
| Yes | 6,485 (22.6%) | 5,583 (86.1%) | 902 (13.9%) |  |
| No | 22,232 (77.4%) | 19,874 (89.4%) | 2,358 (10.6%) |  |
| **Dyspareunia, n (%)** |  |  |  | **<0.001** |
| Yes | 2,539 (8.8%) | 2,196 (86.5%) | 343 (13.5%) |  |
| No | 26,178 (91.2%) | 23,261 (88.9%) | 2,917 (11.1%) |  |
| **Post-menopausal, n (%)** |  |  |  | 0.985 |
| Yes | 2,425 (8.4%) | 2,150 (88.7%) | 275 (11.3%) |  |
| No | 26,292 (91.6%) | 23,307 (88.7%) | 2,985 (11.4%) |  |
| **Dysmenorrhea, n (%)** |  |  |  | **<0.001** |
| Yes | 5,332 (18.6%) | 4,580 (85.9%) | 752 (14.1%) |  |
| No | 23,385 (81.4%) | 20,877 (89.3%) | 2,508 (10.7%) |  |
| **Anxiety Disorders <sup>c</sup>, n (%)** |  |  |  | **0.014** |
| Yes | 18,004 (62.7%) | 15,896 (88.3%) | 2,108 (11.7%) |  |
| No | 10,713 (37.3%) | 9,561 (89.3%) | 1,152 (10.8%) |  |
| **Postpartum depression, n (%)** |  |  |  | 0.703 |
| Yes | 273 (1.0%) | 244 (89.4%) | 29 (10.6%) |  |
| No | 28,444 (99.0%) | 25,213 (88.6%) | 3,231 (11.4%) |  |
| **History of Military Sexual Trauma, n (%)** |  |  |  | **0.041** |
| Yes | 10,981 (38.2%) | 9,681 (88.2%) | 1,300 (11.8%) |  |
| No | 17,736 (61.8%) | 15,776 (88.9%) | 1,960 (11.1%) |  |

<sup>a</sup> Statistical significance was determined by Chi-Square or T-test. Significant values (*p-value* < 0.05) are bolded.
<sup>b</sup> Data for this variable may include instances of missingness that could not be fully accounted for in the analysis
<sup>c</sup> Anxiety disorders included panic disorder (episodic paroxysmal anxiety), generalized anxiety disorder, other specific anxiety disorders, anxiety disorder (unspecified), and other mixed anxiety disorders.

### Patient Characteristics

We found minimal differences in patients’ sociodemographic information among those who received pharmaceutical pain management for their procedure compared to those who received none. For both the pain medication and no pain medication cohorts, average age was around 39 years (SD: 7.6), and roughly 33% had third-party insurance. Slightly higher proportions of Native Hawaiian or Pacific Islanders (13.2%) and Black or African Americans (12.8%) were prescribed pain medication compared to Non-Hispanic Whites (10.6%) and American Indian or Alaskan Natives (8.3%; p<0.001). For marital status, 13.8% of patients who self-identified as “Single” were prescribed pain medication while 10.1% of those self-identified as “Married” were prescribed (p<0.001; **Table 1**)

When analyzing history of childbirths, we found a higher proportion of nulliparous patients prescribed pain medication compared to parous/multiparous patients (11.4% vs 8.9%; p<0.001). Assessing comorbidities revealed that CCI scores were low for both groups, although slightly higher among those not prescribed pain medication (average [SD]: 0.58 [1.13] vs 0.55 [1.05]). Analysis of specific related conditions demonstrated statically significant results for those receiving pain medication and being diagnosed or not (respectively) with the following conditions: anxiety disorders (11.7% vs. 10.8%; p=0.014), dysmenorrhea (14.1% vs. 10.7%; p<0.001), dyspareunia (13.5% vs 11.1%; p<0.001), chronic pelvic pain (13.9% vs 10.6%; p<0.001), and those with a history of MST (11.8% vs. 11.1%; p=0.041; **Table 1**).

### IUD Insertion Procedures and Pharmaceutical Pain Medication Types

Out of the 28,717 IUD insertion procedures captured, NSAIDs were the most commonly prescribed pharmaceutical category (8.3%; **Table 2**), followed by prostaglandins (1.6%), opioid analgesics (0.6%), combination/other (0.6%), and lidocaine (0.2%). The most common clinic location for IUD insertions was gynecology clinics (88.5%), followed by Comprehensive Women’s and Gender Diverse Primary Care Clinic (8.5%). The majority of procedures were performed at a high complexity facility (91.7%) compared to low complexity facilities (3.4%). However, there was relatively small variation in procedure locations where no pain medication was prescribed, ranging from 88.5% at high complexity facilities compared to 92.1% among procedures performed at medium complexity facilities (**Table 2**).

**Table 2:**
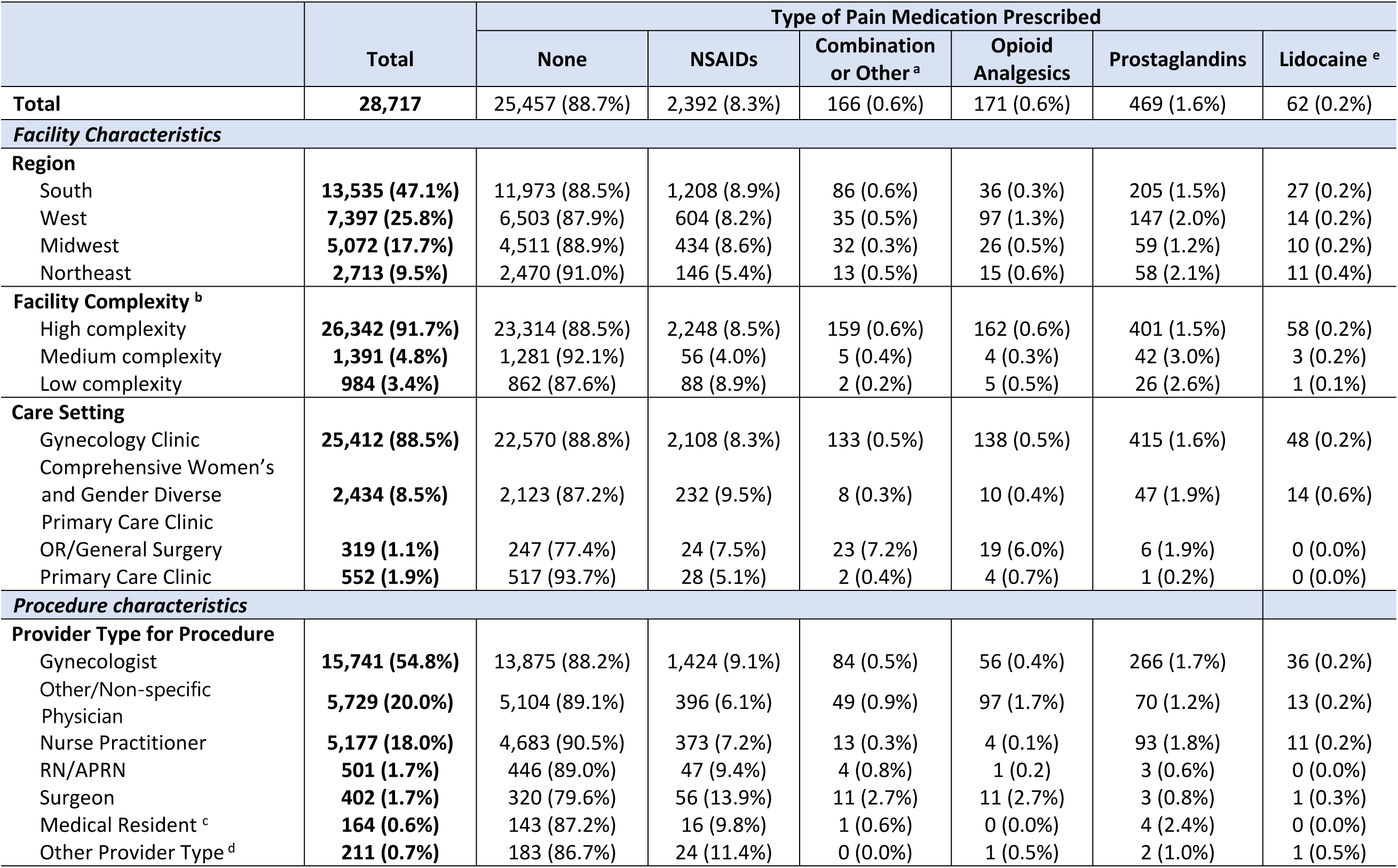
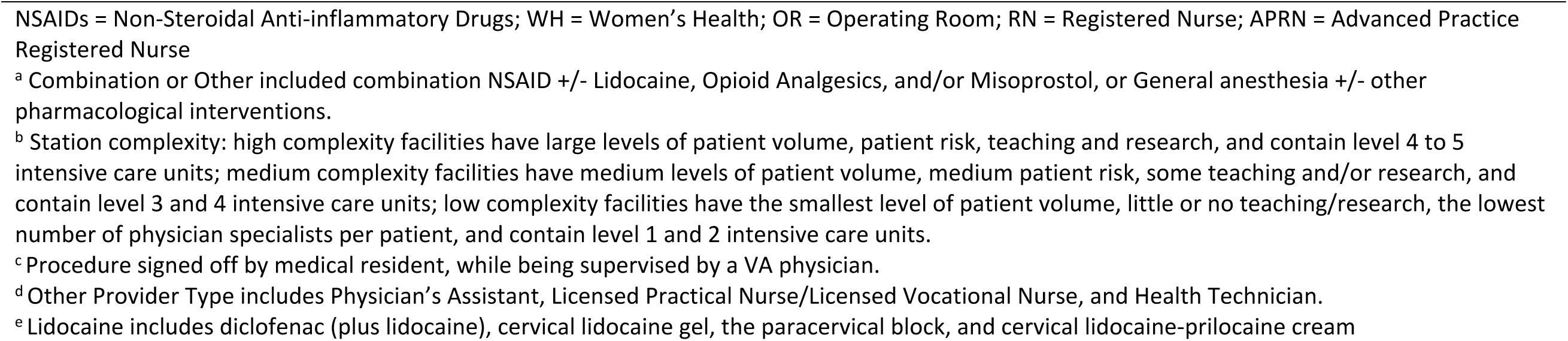
VA clinic region, care setting, primary provider characteristics, and medication types for intrauterine device insertion procedures performed (row frequencies, n (%)).

Procedures were most commonly performed by gynecologists (54.8%), other/non-specific physicians (20.0%), and nurse practitioners (18.0%). There was little variation among provider types in instances where no pain medication was prescribed, ranging from 87.2% among medical residents to 90.5% among nurse practitioners. There was a higher proportion of pain medication prescribed for patients receiving their IUD within the categorical area of the operating room (OR)/general surgery (22.6%), with 7.5% of these patients prescribed an NSAID and 6.0% prescribed an opioid analgesic (**Table 2**).

Among individual medications, ibuprofen was the most frequently prescribed pain intervention as a stand-alone treatment (6.1%) and was also commonly prescribed in combination with other pain intervention modalities (**Table 3**). Additionally, we found that the cervical ripening agent, misoprostol, was prescribed among 1.6% out of the total procedures performed, and ketorolac and naproxen were prescribed among 1.6% and 0.5% of procedures, respectively. Additional information on individual medications and medication combinations per clinic location can be found in **Table 3**.

**Table 3:**
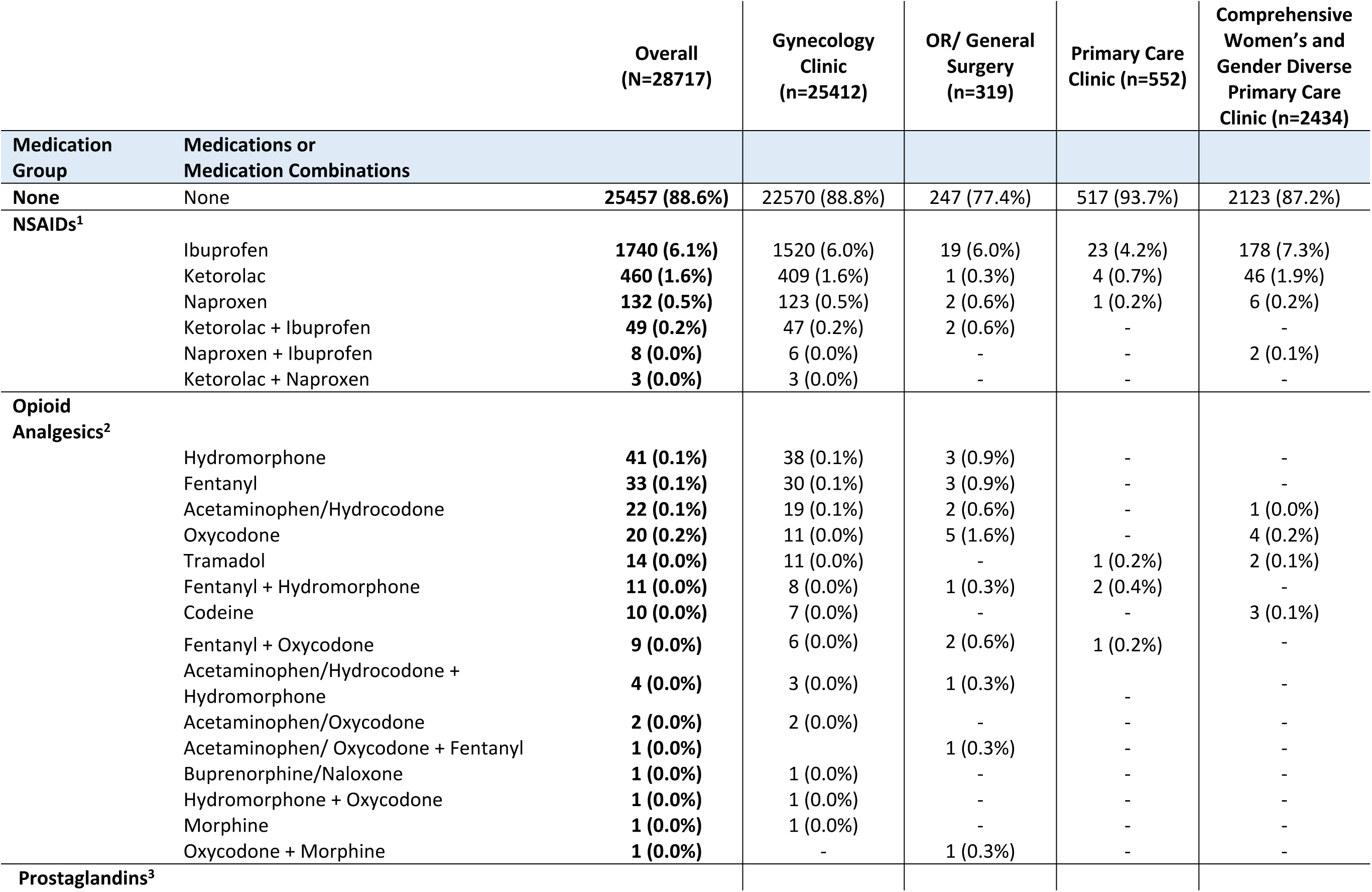
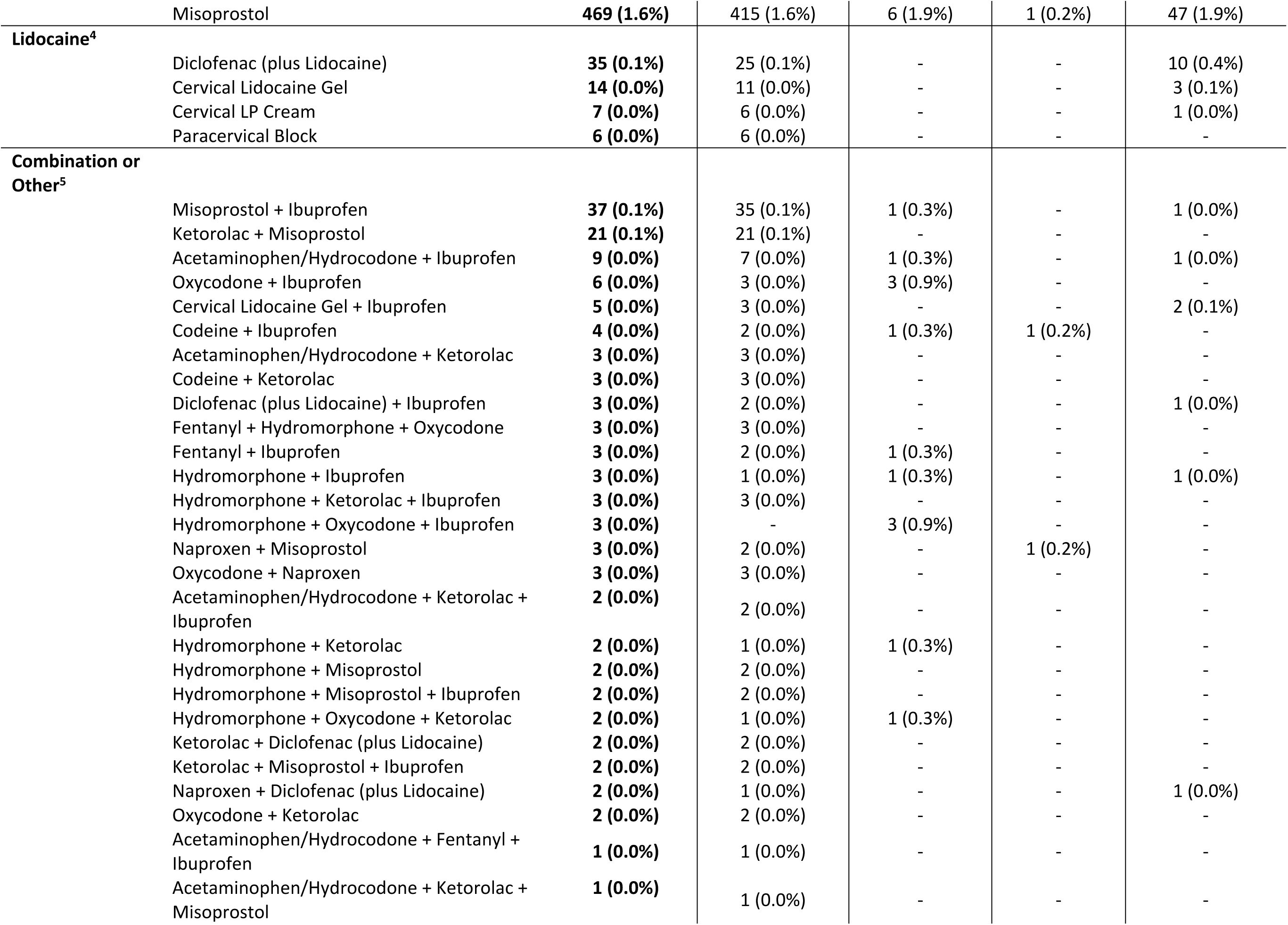
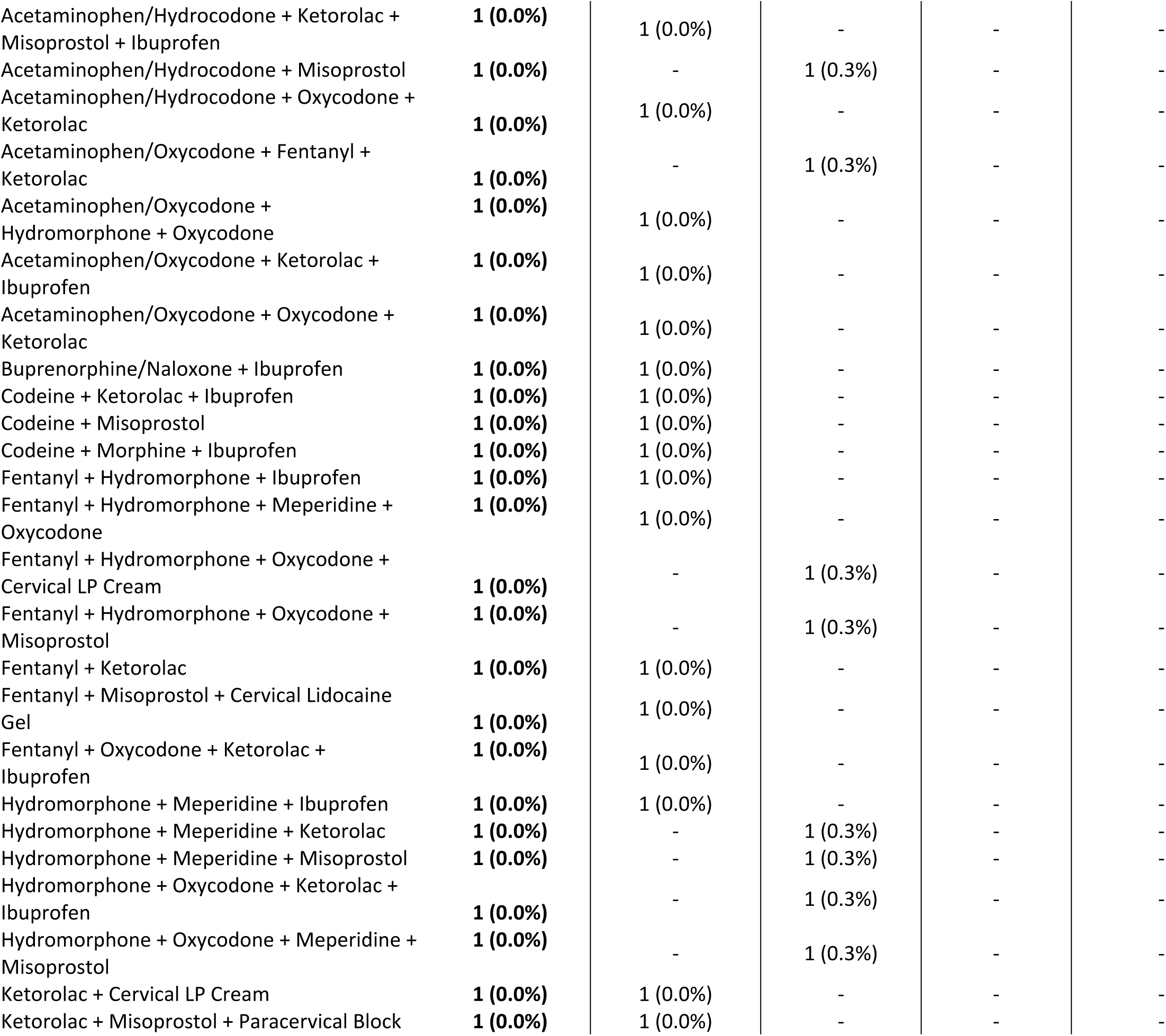
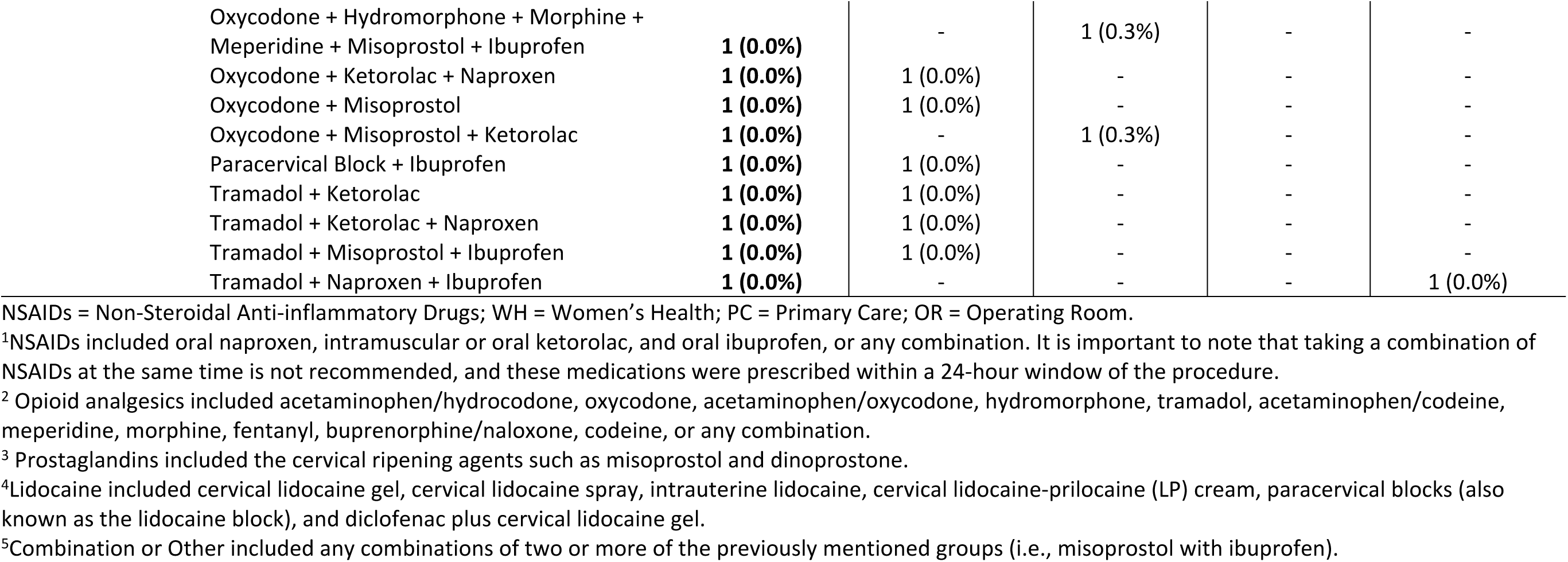
Individual Medications and Medication Combinations Prescribed for Intrauterine Device Insertion Procedure by VA Care Setting.

|  |  | Overall<br>(N=28717) | Gynecology<br>Clinic<br>(n=25412) | OR/ General<br>Surgery<br>(n=319) | Primary Care<br>Clinic (n=552) | Comprehensive<br>Women's and<br>Gender Diverse<br>Primary Care<br>Clinic (n=2434) |
| --- | --- | --- | --- | --- | --- | --- |
| Medication<br>Group | Medications or<br>Medication Combinations |  |  |  |  |  |
| None | None | 25457 (88.6%) | 22570 (88.8%) | 247 (77.4%) | 517 (93.7%) | 2123 (87.2%) |
| **NSAIDs<sup>1</sup>** |  |  |  |  |  |  |
|  | Ibuprofen | 1740 (6.1%) | 1520 (6.0%) | 19 (6.0%) | 23 (4.2%) | 178 (7.3%) |
|  | Ketorolac | 460 (1.6%) | 409 (1.6%) | 1 (0.3%) | 4 (0.7%) | 46 (1.9%) |
|  | Naproxen | 132 (0.5%) | 123 (0.5%) | 2 (0.6%) | 1 (0.2%) | 6 (0.2%) |
|  | Ketorolac + Ibuprofen | 49 (0.2%) | 47 (0.2%) | 2 (0.6%) | - | - |
|  | Naproxen + Ibuprofen | 8 (0.0%) | 6 (0.0%) | - | - | 2 (0.1%) |
|  | Ketorolac + Naproxen | 3 (0.0%) | 3 (0.0%) | - | - | - |
| **Opioid**<br>**Analgesics<sup>2</sup>** |  |  |  |  |  |  |
|  | Hydromorphone | 41 (0.1%) | 38 (0.1%) | 3 (0.9%) | - | - |
|  | Fentanyl | 33 (0.1%) | 30 (0.1%) | 3 (0.9%) | - | - |
|  | Acetaminophen/Hydrocodone | 22 (0.1%) | 19 (0.1%) | 2 (0.6%) | - | 1 (0.0%) |
|  | Oxycodone | 20 (0.2%) | 11 (0.0%) | 5 (1.6%) | - | 4 (0.2%) |
|  | Tramadol | 14 (0.0%) | 11 (0.0%) | - | 1 (0.2%) | 2 (0.1%) |
|  | Fentanyl + Hydromorphone | 11 (0.0%) | 8 (0.0%) | 1 (0.3%) | 2 (0.4%) | - |
|  | Codeine | 10 (0.0%) | 7 (0.0%) | - | - | 3 (0.1%) |
|  | Fentanyl + Oxycodone | 9 (0.0%) | 6 (0.0%) | 2 (0.6%) | 1 (0.2%) | - |
|  | Acetaminophen/Hydrocodone +<br>Hydromorphone | 4 (0.0%) | 3 (0.0%) | 1 (0.3%) | - | - |
|  | Acetaminophen/Oxycodone | 2 (0.0%) | 2 (0.0%) | - | - | - |
|  | Acetaminophen/ Oxycodone + Fentanyl | 1 (0.0%) | - | 1 (0.3%) | - | - |
|  | Buprenorphine/Naloxone | 1 (0.0%) | 1 (0.0%) | - | - | - |
|  | Hydromorphone + Oxycodone | 1 (0.0%) | 1 (0.0%) | - | - | - |
|  | Morphine | 1 (0.0%) | 1 (0.0%) | - | - | - |
|  | Oxycodone + Morphine | 1 (0.0%) | - | 1 (0.3%) | - | - |
| **Prostaglandins<sup>3</sup>** |  |  |  |  |  |  |
|  | Misoprostol | 469 (1.6%) | 415 (1.6%) | 6 (1.9%) | 1 (0.2%) | 47 (1.9%) |
| **Lidocaine<sup>4</sup>** |  |  |  |  |  |  |
|  | Diclofenac (plus Lidocaine) | 35 (0.1%) | 25 (0.1%) | - | - | 10 (0.4%) |
|  | Cervical Lidocaine Gel | 14 (0.0%) | 11 (0.0%) | - | - | 3 (0.1%) |
|  | Cervical LP Cream | 7 (0.0%) | 6 (0.0%) | - | - | 1 (0.0%) |
|  | Paracervical Block | 6 (0.0%) | 6 (0.0%) | - | - | - |
| **Combination or Other<sup>5</sup>** |  |  |  |  |  |  |
|  | Misoprostol + Ibuprofen | 37 (0.1%) | 35 (0.1%) | 1 (0.3%) | - | 1 (0.0%) |
|  | Ketorolac + Misoprostol | 21 (0.1%) | 21 (0.1%) | - | - | - |
|  | Acetaminophen/Hydrocodone + Ibuprofen | 9 (0.0%) | 7 (0.0%) | 1 (0.3%) | - | 1 (0.0%) |
|  | Oxycodone + Ibuprofen | 6 (0.0%) | 3 (0.0%) | 3 (0.9%) | - | - |
|  | Cervical Lidocaine Gel + Ibuprofen | 5 (0.0%) | 3 (0.0%) | - | - | 2 (0.1%) |
|  | Codeine + Ibuprofen | 4 (0.0%) | 2 (0.0%) | 1 (0.3%) | 1 (0.2%) | - |
|  | Acetaminophen/Hydrocodone + Ketorolac | 3 (0.0%) | 3 (0.0%) | - | - | - |
|  | Codeine + Ketorolac | 3 (0.0%) | 3 (0.0%) | - | - | - |
|  | Diclofenac (plus Lidocaine) + Ibuprofen | 3 (0.0%) | 2 (0.0%) | - | - | 1 (0.0%) |
|  | Fentanyl + Hydromorphone + Oxycodone | 3 (0.0%) | 3 (0.0%) | - | - | - |
|  | Fentanyl + Ibuprofen | 3 (0.0%) | 2 (0.0%) | 1 (0.3%) | - | - |
|  | Hydromorphone + Ibuprofen | 3 (0.0%) | 1 (0.0%) | 1 (0.3%) | - | 1 (0.0%) |
|  | Hydromorphone + Ketorolac + Ibuprofen | 3 (0.0%) | 3 (0.0%) | - | - | - |
|  | Hydromorphone + Oxycodone + Ibuprofen | 3 (0.0%) | - | 3 (0.9%) | - | - |
|  | Naproxen + Misoprostol | 3 (0.0%) | 2 (0.0%) | - | 1 (0.2%) | - |
|  | Oxycodone + Naproxen | 3 (0.0%) | 3 (0.0%) | - | - | - |
|  | Acetaminophen/Hydrocodone + Ketorolac + Ibuprofen | 2 (0.0%) | 2 (0.0%) | - | - | - |
|  | Hydromorphone + Ketorolac | 2 (0.0%) | 1 (0.0%) | 1 (0.3%) | - | - |
|  | Hydromorphone + Misoprostol | 2 (0.0%) | 2 (0.0%) | - | - | - |
|  | Hydromorphone + Misoprostol + Ibuprofen | 2 (0.0%) | 2 (0.0%) | - | - | - |
|  | Hydromorphone + Oxycodone + Ketorolac | 2 (0.0%) | 1 (0.0%) | 1 (0.3%) | - | - |
|  | Ketorolac + Diclofenac (plus Lidocaine) | 2 (0.0%) | 2 (0.0%) | - | - | - |
|  | Ketorolac + Misoprostol + Ibuprofen | 2 (0.0%) | 2 (0.0%) | - | - | - |
|  | Naproxen + Diclofenac (plus Lidocaine) | 2 (0.0%) | 1 (0.0%) | - | - | 1 (0.0%) |
|  | Oxycodone + Ketorolac | 2 (0.0%) | 2 (0.0%) | - | - | - |
|  | Acetaminophen/Hydrocodone + Fentanyl + Ibuprofen | 1 (0.0%) | 1 (0.0%) | - | - | - |
|  | Acetaminophen/Hydrocodone + Ketorolac + Misoprostol | 1 (0.0%) | 1 (0.0%) | - | - | - |

|  |  |  |  |  |  |
| --- | --- | --- | --- | --- | --- |
| Acetaminophen/Hydrocodone + Ketorolac + Misoprostol + Ibuprofen | **1 (0.0%)** | 1 (0.0%) | - | - | - |
| Acetaminophen/Hydrocodone + Misoprostol | **1 (0.0%)** | - | 1 (0.3%) | - | - |
| Acetaminophen/Hydrocodone + Oxycodone + Ketorolac | **1 (0.0%)** | 1 (0.0%) | - | - | - |
| Acetaminophen/Oxycodone + Fentanyl + Ketorolac | **1 (0.0%)** | - | 1 (0.3%) | - | - |
| Acetaminophen/Oxycodone + Hydromorphone + Oxycodone | **1 (0.0%)** | 1 (0.0%) | - | - | - |
| Acetaminophen/Oxycodone + Ketorolac + Ibuprofen | **1 (0.0%)** | 1 (0.0%) | - | - | - |
| Acetaminophen/Oxycodone + Oxycodone + Ketorolac | **1 (0.0%)** | 1 (0.0%) | - | - | - |
| Buprenorphine/Naloxone + Ibuprofen | **1 (0.0%)** | 1 (0.0%) | - | - | - |
| Codeine + Ketorolac + Ibuprofen | **1 (0.0%)** | 1 (0.0%) | - | - | - |
| Codeine + Misoprostol | **1 (0.0%)** | 1 (0.0%) | - | - | - |
| Codeine + Morphine + Ibuprofen | **1 (0.0%)** | 1 (0.0%) | - | - | - |
| Fentanyl + Hydromorphone + Ibuprofen | **1 (0.0%)** | 1 (0.0%) | - | - | - |
| Fentanyl + Hydromorphone + Meperidine + Oxycodone | **1 (0.0%)** | 1 (0.0%) | - | - | - |
| Fentanyl + Hydromorphone + Oxycodone + Cervical LP Cream | **1 (0.0%)** | - | 1 (0.3%) | - | - |
| Fentanyl + Hydromorphone + Oxycodone + Misoprostol | **1 (0.0%)** | - | 1 (0.3%) | - | - |
| Fentanyl + Ketorolac | **1 (0.0%)** | 1 (0.0%) | - | - | - |
| Fentanyl + Misoprostol + Cervical Lidocaine Gel | **1 (0.0%)** | 1 (0.0%) | - | - | - |
| Fentanyl + Oxycodone + Ketorolac + Ibuprofen | **1 (0.0%)** | 1 (0.0%) | - | - | - |
| Hydromorphone + Meperidine + Ibuprofen | **1 (0.0%)** | 1 (0.0%) | - | - | - |
| Hydromorphone + Meperidine + Ketorolac | **1 (0.0%)** | - | 1 (0.3%) | - | - |
| Hydromorphone + Meperidine + Misoprostol | **1 (0.0%)** | - | 1 (0.3%) | - | - |
| Hydromorphone + Oxycodone + Ketorolac + Ibuprofen | **1 (0.0%)** | - | 1 (0.3%) | - | - |
| Hydromorphone + Oxycodone + Meperidine + Misoprostol | **1 (0.0%)** | - | 1 (0.3%) | - | - |
| Ketorolac + Cervical LP Cream | **1 (0.0%)** | 1 (0.0%) | - | - | - |
| Ketorolac + Misoprostol + Paracervical Block | **1 (0.0%)** | 1 (0.0%) | - | - | - |
| Oxycodone + Hydromorphone + Morphine +<br>Meperidine + Misoprostol + Ibuprofen | **1 (0.0%)** | - | 1 (0.3%) | - | - |
| Oxycodone + Ketorolac + Naproxen | **1 (0.0%)** | 1 (0.0%) | - | - | - |
| Oxycodone + Misoprostol | **1 (0.0%)** | 1 (0.0%) | - | - | - |
| Oxycodone + Misoprostol + Ketorolac | **1 (0.0%)** | - | 1 (0.3%) | - | - |
| Paracervical Block + Ibuprofen | **1 (0.0%)** | 1 (0.0%) | - | - | - |
| Tramadol + Ketorolac | **1 (0.0%)** | 1 (0.0%) | - | - | - |
| Tramadol + Ketorolac + Naproxen | **1 (0.0%)** | 1 (0.0%) | - | - | - |
| Tramadol + Misoprostol + Ibuprofen | **1 (0.0%)** | 1 (0.0%) | - | - | - |
| Tramadol + Naproxen + Ibuprofen | **1 (0.0%)** | - | - | - | 1 (0.0%) |

NSAIDs = Non-Steroidal Anti-inflammatory Drugs; WH = Women's Health; PC = Primary Care; OR = Operating Room.
<sup>1</sup>NSAIDs included oral naproxen, intramuscular or oral ketorolac, and oral ibuprofen, or any combination. It is important to note that taking a combination of NSAIDs at the same time is not recommended, and these medications were prescribed within a 24-hour window of the procedure.
<sup>2</sup>Opioid analgesics included acetaminophen/hydrocodone, oxycodone, acetaminophen/oxycodone, hydromorphone, tramadol, acetaminophen/codeine, meperidine, morphine, fentanyl, buprenorphine/naloxone, codeine, or any combination.
<sup>3</sup>Prostaglandins included the cervical ripening agents such as misoprostol and dinoprostone.
<sup>4</sup>Lidocaine included cervical lidocaine gel, cervical lidocaine spray, intrauterine lidocaine, cervical lidocaine-prilocaine (LP) cream, paracervical blocks (also known as the lidocaine block), and diclofenac plus cervical lidocaine gel.
<sup>5</sup>Combination or Other included any combinations of two or more of the previously mentioned groups (i.e., misoprostol with ibuprofen).

### Geographic variation in pain treatment

Our analysis revealed regional variations throughout the US regarding the prescription of pain medication for IUD insertions, with the proportion of those prescribed any form of pain medication varying from 9.0% in the Northeast to 12.1% in the West (**Table 2**). However, a closer examination at the state level uncovered more significant differences, with the percentage of procedures involving any form of pain medication ranging from 1.0% in Missouri to 32.0% in New Hampshire (**Figure 2, and Appendix 3**). Furthermore, our results indicate that the prescription of pain medications other than ibuprofen for procedures across the US states was notably low, varying from 0.6% in Missouri to 26.7% in the District of Columbia (**Appendix 3**).

**Figure 2:**
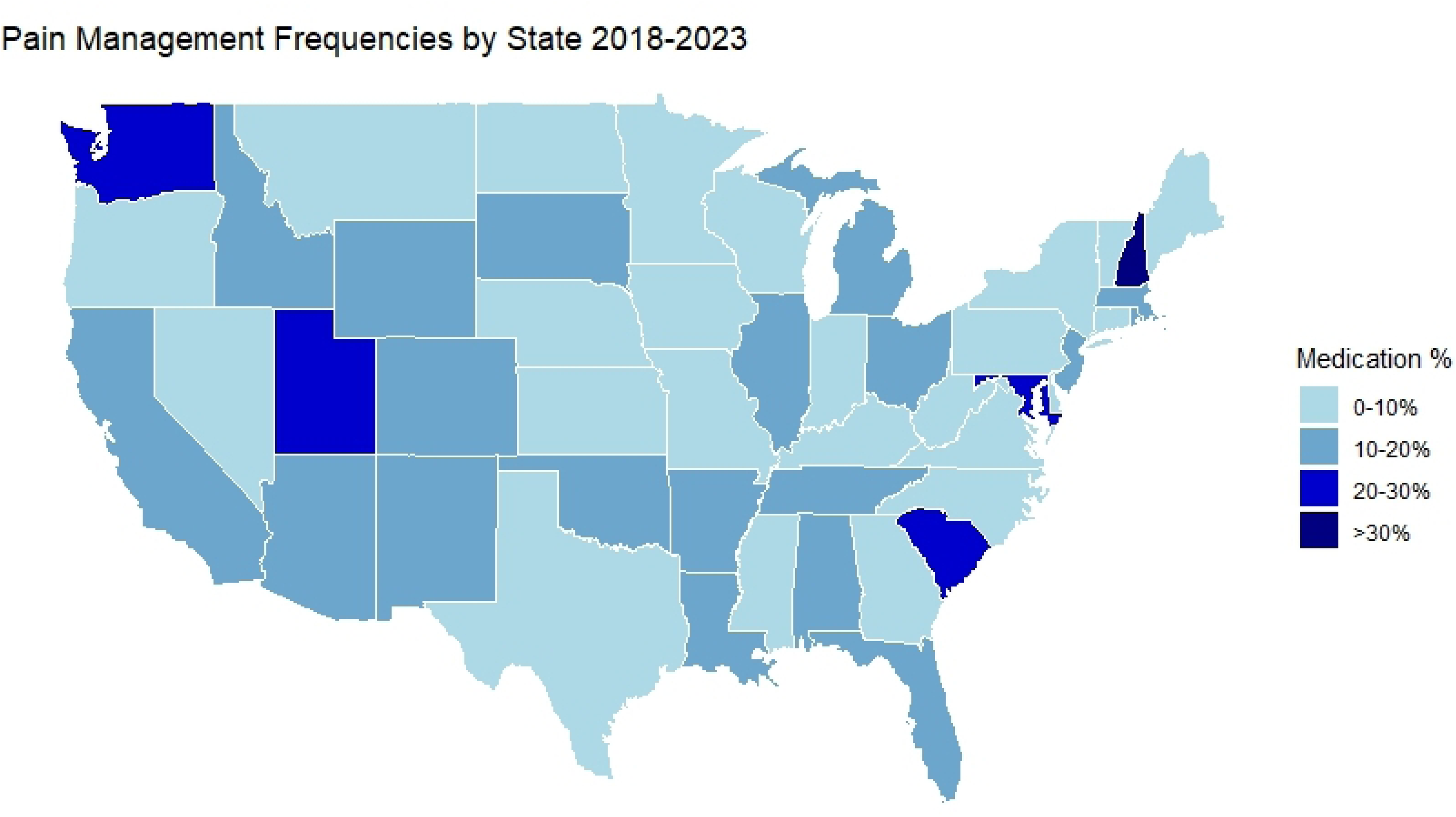
Percentage among the 24,010 IUD insertions performed within VHA prescribed pain medication across the United States, January 1st, 2018, to October 13th, 2023. **F2L:** The blue scale gradient on the United States map above depicts the average proportion of intrauterine device insertion procedures where any pain medication was prescribed within the Veterans Affairs Healthcare System from January 1^st^, 2018, to October 13^th^, 2023, with the darker blues associated with a higher proportion of procedures with prescribed medications. Exact percentages can be found in S2.

## Discussion

IUDs are a common form of long-acting reversible contraceptives. Unfortunately, the many patients report pain during and after placement, with over 75% rating their pain as moderate to severe.^5^ Over our assessment timeframe, only 11.4% of the 28,717 IUD insertions performed at VHA, nationally, had any form of documented pain medication. However, there was a steady increase in the proportion of those provided pain medication options, with an average annual increase of 0.52% between January 1st, 2018, to October 13th, 2023.

There was significant variation between US states in the proportion with patients who received any form of pain medication ranging from 1.0% in Missouri to 32.0% in New Hampshire, even though the majority of the IUD insertion procedures were performed within high complexity facilities (91.7%) with similar available resources.^32^ Depending on location, only 0.6% to 26.7% of procedures were prescribed analgesics other than ibuprofen. While our data could not assess the potential causes of the variation, possible factors include regional practices, provider training, patient preferences, and/or specific protocols within each VHA facility.

Despite multiple studies concluding ibuprofen is ineffective at reducing pain during or after IUD insertion,^3,33,34^ it was the most frequently prescribed intervention for pain. Previous research has shown that naproxen is the only NSAID shown to be effective in decreasing pain during IUD insertion.^35,36^ Interestingly, our results show naproxen, as a stand-alone treatment, was only prescribed 0.5% of the time. Other options, like prostaglandins, which are often used to induce cervical ripening in vaginal delivery, were at one time thought to be effective in helping ease IUD placement and, therefore, any associated pain.^37^ However, more recently published results have found prostaglandins to be ineffective in easing the IUD insertion process and associated pain reduction.^37,38^ Only 1.6% of procedures captured in this assessment were prescribed prostaglandins.

A small number of prescribed opioid analgesics were prescribed, potentially for patients with a more complicated health history than our data was unable to uncover (i.e., previously embedded IUD, scarring, etc.). To our knowledge, most opioids have not yet been tested for their effectiveness in reducing pain during or after an IUD procedure with the exception of tramadol, which has been shown to be effective in reducing pain with IUD placement.^33,39^ In this assessment, tramadol was prescribed in less than 1% of procedures, often in combination with other prescribed pain interventions.

Various formulations of lidocaine have been shown to be effective in reducing pain associated with IUD insertions. For example, cervical lidocaine 2% gel (either with or without the combination of diclofenac) has shown some effectiveness for reducing pain associated with IUD placement.^40,41^ Cervical lidocaine 4% gel, LP (lidocaine; prilocaine) cream, and the paracervical block with lidocaine have all been shown to be effective either with tenaculum placement or after the procedure.^14,40^ LP cream alone has also been shown to reduce pain of tenaculum placement by 24% and pain from IUD insertion by 28%.^42^ Yet despite this evidence, only 0.2% of the procedures captured in our sample were prescribed any form lidocaine. Additionally, paracervical blocks have been shown to be effective in reducing overall pain associated with this procedure. ^42^ However, we found only 6 occurrences (0.0%) of the paracervical block being prescribed as a standalone pain intervention for IUD insertions within our sample, and only 8 occurrences within the dataset in its entirety.

Some patients may experience more pain or discomfort than others during IUD insertion depending on their medical history. Previous studies have shown that patients with a history of chronic pelvic pain, sexual pain, dysmenorrhea, or painful periods, as well as those who are post-partum, postmenopausal, have a history of sexual trauma, and those who suffer from anxiety, are more likely to experience greater pain with IUD insertions.^43^ We found 62.7% of our cohort were diagnosed with anxiety disorders, 38.2% had a history of MST, and 22.6% had a history of chronic pelvic pain. Additionally, nulliparous patients have been found to report significantly higher pain when undergoing IUD insertions compared to parous patients, ^44^ which comprised 94.4% of our sample. As such, it is important to assess the efficacy of different options in our specific VHA patient population, as results of work outside of our patient population may not necessary be generalizable. Furthermore, assessing these types of factors and bridging gaps in care are an important part of informing a comprehensive care strategy.

Nonetheless, our data demonstrates a significant lack of prescribed pain interventions for IUD insertions, and when initiated, VHA providers were disproportionally prescribing medications that have not been shown to be effective in reducing pain during or after the IUD insertion. Furthermore, there was a significant geographic variation in pain treatment practices, which suggests a lack of a comprehensively implemented strategy across the VHA. Other countries have systematically taken steps to address analgesic options for pain related to IUD placements, and understanding these efforts may provide policy insights. For example, in 2020, clinical guidelines with recommended pain interventions were adopted in Canada and reflected in an official statement by the Society of Obstetricians and Gynecologists of Canada in 2023.^43,45^ These guidelines strongly recommend that healthcare providers counsel patients on the various pain management options available before the insertion procedure, taking into account the patient’s medical history and the specific techniques used, as these factors can influence the effectiveness of the pain intervention. Additionally, a public petition in the United Kingdom amassed over 35,000 signatures advocating for better pain relief for IUD insertions, leading the Faculty of Sexual and Reproductive Healthcare and the Royal College of Obstetricians and Gynecologists to release a statement updating their guidance to members; with the president, Dr Edward Morris, stating, “…*We believe that unbearable pain during any gynecological procedure is unacceptable and all specialists working in women’s health; specialist nurses, GPs and gynecologists need to listen and take account of what is being said*.”^46^

There could be several reasons for the lack of an apparent comprehensive approach in our population. For example, one reported potential reason could be that some providers perceive pain to be less than the patient experiences during gynecological procedures.^48^ In further support of this, a 2015 study found that the mean patient maximum pain during the IUD insertion was 64.8 millimeters (mm) on a 100mm visual analogue scale (where 0mm represents “no pain” and 100mm represents “the worst imaginable pain”) compared to only 35.3mm when rated by the provider.^47^ In addition, a 2018 meta-analysis on gender bias in healthcare suggests that the lack of pain medication may be due to a healthcare system’s failure to legitimize women’s pain and revealed that a women’s pain is more likely to be described as “hysterical” or “sensitive.”^48^ By initiating standardized and data informed clinical guidelines for providers to counsel patients on what to expect during IUD placement, including effective pain control options, we believe that VHA will not only provide better care, but also strengthen the trust between Veterans and their providers.

Our VA population may face unique Veteran-related health challenges, which may limit the generalizability of our assessment to non-VA settings. This assessment is limited as it is a retrospective analysis of routinely collected clinical data demonstrating population level associations rather than causations. However, this retrospective design also provides valuable real-world insights about nationwide clinical trends and practices that may be challenging to achieve in a prospective trial due to enrollment size and the Hawthorne effect.^49^ Although we queried both structured and unstructured data fields to capture all analgesic options, we may have missed medications purchased over the counter (OTC) or prescribed outside of the VA. Despite this, because VA practices purposeful adverse patient selection, many Veterans rely on VA for OTC medications, offering a broader scope of care and a more complete record of outpatient analgesic use. However, Veterans with better financial means may not rely on VA for OTC medications, and it is possible that some of these patients did not report their pain medication use to their clinicians, even though medication reconciliation is a national VA system-wide policy for clinical visits, and the VA EHR database is designed to include non-VA prescribed OTC medications. Furthermore, our method of capturing any non-opioid pain medication within 45 days of IUD placement was designed to comprehensively include any potential pain medication associated with the procedure. Although this method increased the sensitivity for detecting prescribed pain medications, it reduced the specificity that the medications prescribed during this time were intended for IUD associated pain. As a result, the rates of pain medication prescription for IUD placement may actually be even lower than we reported, further emphasizing the care gap and need for informed action.

A notable strength of this assessment is the large population-based sample of Veterans, from across the US, utilizing documented clinical data (e.g., diagnoses, medication use, and demographics). In addition, our approach leveraged both structured and unstructured data sources from both inpatient and outpatient sources, to provide a comprehensive assessment of pain prescriptions. Future work to analyze optimal pain strategies for our specific patient population, analyzing potential changes after data informed policies are implemented, as well as understanding patient characteristics that are predictive of being prescribed pain interventions will be of additional value.

The aim of this assessment was to evaluate the current state of prescribed pain intervention for our Veteran population undergoing IUD insertion procedures at a national level. This evaluation is a crucial step towards improving pain management practices. Our findings will inform the development and adoption of evidence-based guidelines and the standardization of pain management protocols. Enhancing provider education is paramount to ensure healthcare professionals are well-informed about effective pain control options. Additionally, promoting patient-centered care models that respect the unique needs of Veterans who are AFAB and integrating patient feedback into care processes will be essential. These measures will ensure that pain management during IUD insertion procedures is both effective and tailored to individual patient needs. After implementing these guidelines and educational interventions, we plan to reassess the outcomes to continuously improve the quality of care provided.

## Conclusion

Although there has been a steady increase in the use of prescribed pain treatment for IUD procedures in the VHA, the overall rate of prescribed pain interventions for IUD procedures remains low. Furthermore, the most commonly prescribed analgesic medications for IUD insertions in VHA are different than what the current literature has shown to be effective treatments. The intent of this assessment is that it will contribute to data informed interventions, policies, education, and processes that promote optimal standardized pain medications for patients undergoing IUD insertion procedures within the VHA.

## Data Availability

Due to US Department of Veterans Affairs (VA) regulations and ethics agreements, the data utilized for this assessment are not permitted to leave the VA firewall without a Data Use Agreement. However, VA data can be made available to researchers with an approved VA authorized protocol.

## References

1 MacMillan C. Yale Medicine [Internet]. Intrauterine Devices (IUDs): What Women Need to Know; 2023 Oct 4. Available at: https://www.yalemedicine.org/news/intrauterine-devices-iud (Accessed December 5, 2023)

2 Jennie Yoost (2014) Understanding benefits and addressing misperceptions and barriers to intrauterine device access among populations in the United States, Patient Preference and Adherence, 8: 947–957, DOI: 10.2147/PPA.S45710

3 Hubacher D, Reyes V, Lillo S, et al. Pain from copper intrauterine device insertion: randomized trial of prophylactic ibuprofen, Am J Obstet Gynecol, 2006, 195; 1272–1277, DOI: 10.1016/j.ajog.2006.08.022

4 Allen RH, Bartz D, Grimes DA, et al. Interventions for pain with intrauterine device insertion, Cochrane Database Syst Rev, 2009, CD007373

5 Hall AM, Kutler BA. Intrauterine contraception in nulliparous women: a prospective survey. J Fam Plann Reprod Health Care. 2016 Jan;42(1):36–42. doi: 10.1136/jfprhc-2014-101046. Epub 2015 Apr 8. PMID: 25854550; PMCID: PMC4717389.

6 Nguyen L, Lamarche L, Lennox R, et al. Strategies to Mitigate Anxiety and Pain in Intrauterine Device Insertion: A Systematic Review. J Obstet Gynaecol Can. 2020 Sep;42(9):1138–1146.e2. doi: 10.1016/j.jogc.2019.09.014. Epub 2019 Dec 25. PMID: 31882291.

7 Lopez LM, Bernholc A, Zeng Y, et al. Interventions for pain with intrauterine device insertion. Cochrane Database of Systematic Reviews 2015, Issue 7. Art. No.: CD007373. DOI: 10.1002/14651858.CD007373.pub3. Accessed 05 December 2023.

8 Gemzell-Danielsson K., Mansour D, Fiala C, et al. Management of pain associated with the insertion of intrauterine contraceptives, Human Reproduction Update, Volume 19, Issue 4, July/August 2013, Pages 419–427, 10.1093/humupd/dmt022jon

9 Faustino R. Perez-Lopez, Samuel J. et al. Uterine or paracervical lidocaine application for pain control during intrauterine contraceptive device insertion: a meta-analysis of randomised controlled trials, The European Journal of Contraception & Reproductive Health Care, 2018. 23:3, 207–217, DOI: 10.1080/13625187.2018.1469124

10 Grimes DA, Hubacher D, Lopez LM, et al. Non-steroidal anti-inflammatory drugs for heavy bleeding or pain associated with intrauterine-device use. Cochrane Database of Systematic Reviews 2006, Issue 4. Art. No.: CD006034. DOI: 10.1002/14651858.CD006034.pub2. Accessed 05 December 2023.

11 Anthoulakis C, Iordanidou E, Vatopoulou A. Pain Perception during Levonorgestrel-releasing Intrauterine Device Insertion in Nulliparous Women: A Systematic Review. J Pediatr Adolesc Gynecol. 2018 Dec;31(6):549–556.e4. doi: 10.1016/j.jpag.2018.05.008. Epub 2018 Jun 8. PMID: 29890206.

12 Gemzell-Danielsson K., Jensen JT., Monteiro I, et al. Interventions for the prevention of pain associated with the placement of intrauterine contraceptives: An updated review. Acta Obstet Gynecol Scand. 2019; 98: 1500–1513. 10.1111/aogs.13662

13 Mody SK, Farala JP, Jimenez B, et al. Paracervical Block for Intrauterine Device Placement Among Nulliparous Women: A Randomized Controlled Trial. Obstet Gynecol. 2018 Sep;132(3):575–582. doi: 10.1097/AOG.0000000000002790. PMID: 30095776; PMCID: PMC6438819.

14 Johnson BA. Insertion and removal of intrauterine devices. Am Fam Physician. 2005 Jan 1;71(1):95–102. PMID: 15663031.

15 Moot G. Service95 [Internet]. Why Is Pain Relief Not Offered for IUD Insertion? - Service95; 2023 Mar 6 [cited 2023 Dec 5]. Available from: https://www.service95.com/why-is-pain-relief-not-offered-for-iud-insertion/.

16 Planned Parenthood | Official Site [Internet]. What’s an IUD insertion like?; 2024 Jan [cited 2024 Mar 20]. Available from: https://www.plannedparenthood.org/learn/birth-control/iud/whats-an-iud-insertion-like

17 Wu J, Trahair E, Happ M, et al. TikTok,# IUD, and user experience with intrauterine devices reported on social media. Obstetrics & Gynecology. 2023 Jan 30;141(1):215–7.

18 Oerman A. Cosmopolitan [Internet]. It’s Time We Talk About IUD Insertion and Pain, No?; 2023 Feb 22 [cited 2023 Dec 5]. Available from: https://www.cosmopolitan.com/health-fitness/a42957255/iud-pain/.

19 Emanuelli A. Chatelaine [Internet]. Getting An IUD Can Really Hurt. Here’s What You Can Do About It; 2023 Oct 19 [cited 2023 Dec 5]. Available from: https://chatelaine.com/health/iud-pain-management/.

20 O’Donohue S. The BMJ [Internet]. The ripples of trauma caused by severe pain during IUD procedures - The BMJ; 2021 Jul 1 [cited 2023 Dec 5]. Available from: https://blogs.bmj.com/bmj/2021/07/20/the-ripples-of-trauma-caused-by-severe-pain-during-iud-procedures/ (Accessed: 1 November 2023).

21 Department of Veterans Affairs [Internet]. Veteran population projections model (VetPop2020). Washington, DC: Department of Veterans Affairs, 2020. Available at: Veteran Population - National Center for Veterans Analysis and Statistics (va.gov). Accessed September 13, 2023.

22 Batuman F, Bean-Mayberry B, Goldzweig C, et al. Health Effects of Military Service on Women Veterans. Department of Veterans Affairs (US), Washington (DC); 2011. PMID: 21796832.

23 US Department of Veterans Affairs, V.H.A. [Internet] Veterans Affairs, Facts and Statistics, 2022. Available at: https://www.womenshealth.va.gov/materials-and-resources/facts-and-statistics.asp (Accessed: 13 September 2023).

24 The White House [Internet]. FACT SHEET: President Biden Issues Executive Order and Announces New Actions to Advance Women’s Health Research and Innovation | The White House 2024. Available at: FACT SHEET: President Biden Issues Executive Order and Announces New Actions to Advance Women’s Health Research and Innovation | The White House (Accessed 21 March 2024).

25 Washington DL. Findings from the National Survey of Women Veterans. Proceedings of the VA HSR&D Cyber Seminar, January 12, 2011, Los Angeles, CA.

26 Department of Veterans Affairs [Internet]. Women’s Health Research. Washington, DC: Department of Veterans Affairs, 2024. Available at: Women’s Health Research (va.gov). Accessed September 13, 2023.

27 Borrero S, Callegari LS, Zhao X, et al. Unintended Pregnancy and Contraceptive Use Among Women Veterans: The ECUUN Study. J Gen Intern Med. 2017 Aug;32(8):900–908. doi: 10.1007/s11606-017-4049-3. Epub 2017 Apr 21. PMID: 28432564; PMCID: PMC5515789.

28 Koenig AF, Borrero S, Zhao X, et al. Factors associated with long-acting reversible contraception use among women Veterans in the ECUUN study. Contraception. 2019 Sep;100(3):234–240. doi: 10.1016/j.contraception.2019.05.010. Epub 2019 May 29. PMID: 31152697; PMCID: PMC6861159.

29 ACOG: Long-Acting Reversible Contraception (LARC) Quick Coding Guide [Internet]. American College of Obstetricians and Gynecologists [cited 2023 Nov 20]. Available from: https://www.acog.org/education-and-events/publications/larc-quick-coding-guide/clinical-scenarios (Accessed 12 March 2024).

30 US Department of Veterans Affairs . Corporate data warehouse (CDW). Available at: www.hsrd.research.va.gov/for_researchers/vinci/cdw.cfm (Accessed 2 January 2024).

31 Centers of Disease Control and Prevention [Internet]. Geographic division or region. Available at: Geographic division or region - Health, United States (cdc.gov) (Accessed: 13 September 2023).

32 National Academies of Sciences, Division of Behavioral, Board on Human-Systems Integration, Division on Engineering, Physical Sciences, Board on Infrastructure, … & Committee on Facilities Staffing Requirements for Veterans Health Administration. (2020). Facilities Staffing Requirements for the Veterans Health Administration Resource Planning and Methodology for the Future.

33 Bednarek PH, Creinin MD, Reeves MF, Cwiak C, Espey E, Jensen JT; Post-Aspiration IUD Randomization (PAIR) Study Trial Group. Prophylactic ibuprofen does not improve pain with IUD insertion: a randomized trial. Contraception. 2015 Mar;91(3):193–7. doi: 10.1016/j.contraception.2014.11.012. Epub 2014 Nov 25. PMID: 25487172.

34. ^34^ Chor J, Bregand-White J, Golobof A, et al. Ibuprofen prophylaxis for levonorgestrel-releasing intrauterine system insertion: a randomized controlled trial. Contraception. 2012 Jun;85(6):558–62. doi: 10.1016/j.contraception.2011.10.015. Epub 2011 Dec 15. PMID: 22176793.

35 Karabayirli S, Ayrim AA, Muslu B. Comparison of the analgesic effects of oral tramadol and naproxen sodium on pain relief during IUD insertion. J Minim Invasive Gynecol. 2012 Sep-Oct;19(5):581–4. doi: 10.1016/j.jmig.2012.04.004. Epub 2012 Jul 4. PMID: 22766124.

36 Edgren RA, Morton CJ. Naproxen sodium for OB/GYN use, with special reference to pain states: a review. International Journal of Fertility. 1986 May-Jun;31(2):135–142. PMID: 2875035.

37 Mansy, Amr Adel. “Does sublingual misoprostol reduce pain and facilitate IUD insertion in women with no previous vaginal delivery? A randomized controlled trial.” Middle East Fertility Society Journal 23.1 (2018): 72–76.

38 Edelman, Alison B., et al. “Effects of prophylactic misoprostol administration prior to intrauterine device insertion in nulliparous women.” Contraception 84.3 (2011): 234–239.

39 Naing K, Shah S. Pain Medications Before IUD Placement. American Family Physician. 2020 Jan 15;101(2):119–20.

40 Akers AY. Pharmacologic approaches to pain management with IUD insertion. Optimizing IUD Delivery for Adolescents and Young Adults: Counseling, Placement, and Management. 2019:111–21.

41 Fouda UM, Eldin NM, Elsetohy KA, et al. Diclofenac plus lidocaine gel for pain relief during intrauterine device insertion. A randomized, double-blinded, placebo-controlled study. Contraception. 2016 Jun 1;93(6):513–8.

42 Whitworth K, Neher J, Safranek S. Effective analgesic options for intrauterine device placement pain. Can Fam Physician. 2020 Aug;66(8):580–581. PMID: 32817031; PMCID: PMC7430802.

43 Ireland, Luu Doan MD, MPH*; Allen, Rebecca H. MD, MPH†. Pain Management for Gynecologic Procedures in the Office. Obstetrical & Gynecological Survey 71(2):p 89–98, February 2016. | DOI: 10.1097/OGX.0000000000000272

44 Lena Marions, Lena Lövkvist, Annika Taube, et al. (2011) Use of the levonorgestrel releasing-intrauterine system in nulliparous women – a non-interventional study in Sweden, The European Journal of Contraception & Reproductive Health Care, 16:2, 126–134, DOI: 10.3109/13625187.2011.558222

45 Statement on Intrauterine Devices, Counselling and Pain Management [Internet]. Ottowa: The Society of Obstetricians and Gynaecologists of Canada; 2022 Dec 15 [cited 2023 Dec 5]. Available at: https://sogc.org/common/Uploaded%20files/Latest%20News/Statement_on_Intrauterine_Devices-E.pdf

46 Faculty of Sexual and Reproductive Healthcare (FSRH) [Internet] FSRH and RCOG press statement on intrauterine device (IUD) insertions and pain relief, 2021. Available at: FSRH and RCOG press statement on intrauterine device (IUD) insertions and pain relief - Faculty of Sexual and Reproductive Healthcare (Accessed: 13 September 2023).

47 Maguire K, Morrell K, Westhoff C, Davis A. Accuracy of providers’ assessment of pain during intrauterine device insertion. Contraception. 2014 Jan;89(1):22–4. doi: 10.1016/j.contraception.2013.09.008. Epub 2013 Sep 21. PMID: 24134898.

48 Samulowitz A, Gremyr I, Eriksson E, et al. ”Brave Men” and “Emotional Women”: A Theory-Guided Literature Review on Gender Bias in Health Care and Gendered Norms towards Patients with Chronic Pain. Pain Res Manag. 2018 Feb 25;2018:6358624. doi: 10.1155/2018/6358624. PMID: 29682130; PMCID: PMC5845507.

49 Sedgwick P, Greenwood N. Understanding the Hawthorne effect. Bmj. 2015 Sep 4;351.

